# A Phase I, Prospective, Randomized, Open-labeled Study to Evaluate the Safety, Tolerability, and Immunogenicity of Booster Dose with MVC-COV1901 or MVC-COV1901(Beta) SARS-CoV-2 Vaccine in Adults

**DOI:** 10.1101/2022.08.29.22279317

**Authors:** Chia En Lien, Ming-Che Liu, Ning-Chi Wang, Luke Tzu-Chi Liu, Chung-Chin Wu, Wei-Hsuan Tang, Wei-Cheng Lian, Kuan-Ying A. Huang, Charles Chen

## Abstract

**Background:** The use of variant-based severe acute respiratory syndrome coronavirus 2 (SARS-CoV-2) vaccine as a booster is being evaluated to overcome reduced neutralisation of variants induced by the original SARS-CoV-2 vaccine and waning protection over time.

**Methods:** This is a phase one, prospective, randomized, and open-labeled trial to study the safety and immunogenicity of a booster dose consisting of a subunit vaccine based on the stabilized prefusion SARS-CoV-2 spike protein, MVC-COV1901 or its Beta version, MVC-COV1901-Beta. One-hundred and seven participants aged ≥18 and <55 years, who received two or three prior doses of MVC-COV1901 vaccines, were enrolled and were to receive a booster dose of either 15 mcg of MVC-COV1901, 15 mcg or 25 mcg of MVC-COV1901-Beta in 1:1:1 ratio. The primary endpoints were the incidences of adverse events and immunogenicity of the booster dose from Visit 2 (the day of the booster) to Visit 5 (four weeks after the booster). Cellular immunity was also investigated with memory B cell (MBC) and T cell assays.

**Findings:** Adverse reactions after either MVC-COV1901 or MVC-COV1901-Beta booster doses after two or three doses of MVC-COV1901 were comparable and mostly mild and transient. At four weeks after the booster dose, participants with two prior doses of MVC-COV1901 exhibited numerically higher levels of neutralising antibodies against SARS-CoV-2 or Beta variant than participants with three prior doses of MVC-COV1901 regardless of the type of booster used. However, compared to 15 mcg of MVC-COV1901, 25 mcg of MVC-COV1901-Beta significantly improved neutralising antibody titre against Beta variant and BA.4/BA.5 Omicron variant pseudoviruses. The booster dose also significantly increased the proportion of spike-specific MBCs, including those of Beta and Omicron variants.

**Interpretation:** MVC-COV1901-Beta can be effectively used as a booster dose against SARS-CoV-2, including the circulating BA.4/BA.5 Omicron variant.

**Funding:** Medigen Vaccine Biologics Corporation

## Introduction

The vaccine efficiency against the SARS-CoV-2 virus is hampered by the circulation of variants resistant to antibody neutralisation imparted by the SARS-CoV-2 vaccine based on the ancestral strain [1, 2]. As of August 2022, WHO has updated its SAR-CoV-2 list of variants of concern (VoC) to consist solely of the Omicron variant and its subvariants [3]. The implication of the Omicron variants as the circulating VoC is that primary series and even booster vaccination with currently available vaccines have shown to perform poorly against the current circulating Omicron variant and its subvariants in general, including the BA.2 and BA.4/BA.5 subvariants [4, 5]. Various Omicron variant-based vaccines have been developed or are in development, including a monovalent mRNA vaccine based on the Omicron variant by Pfizer/BNT is in the midst of a phase 2/3 clinical trial that claimed to have demonstrated effective neutralisation against the BA.1 subvariant and to a lesser extent, the BA.4 and BA.5 subvariants [6]. Recently, citing a strong immune response generated against the ancestral virus and the BA.1 Omicron variant, as well as a good immune response against the BA.4 and BA.5 subvariants, the United Kingdom became the first country in the world to approve a bivalent COVID-19 booster vaccine for adults (Moderna bivalent mRNA vaccine based on the ancestral SARS-CoV-2 and the Omicron variant) [7].

MVC-COV1901 is a subunit COVID-19 vaccine based on the stable prefusion spike protein S-2P of the ancestral SARS-CoV-2 and adjuvanted with CpG 1018 and aluminum hydroxide [8]. We have previously shown that three doses of MVC-COV1901 can improve neutralising antibody response against live SARS-CoV-2 and Omicron variant pseudovirus [9]. We first demonstrated the ability of the Beta variant version of S-2P to protect hamsters from the Delta variant challenge and have also shown improved neutralising antibody levels against a broad spectrum of variants, including the current Omicron variant [10]. In the current study, two groups of participants who have received either two or three doses of MVC-COV1901 were administered with a booster dose of either original MVC-COV1901 or MVC-COV1901 based on the Beta variant in two different dose levels (MVC-COV1901-Beta). Reacogenicity and immunogenicity against the original SARS-COV-2 and beta variant were examined after the booster dose. To investigate the levels of neutralising antibodies against the Omicron subvariants, we have also conducted a pseudovirus neutralisation assay with the Omicron BA.4/BA.5 pseudovirus.

## Methods

### Study design and participants

This was a prospective, randomized, open-labeled phase I study to evaluate the safety, tolerability, and immunogenicity of booster dose with MVC-COV1901 or MVC-COV1901-Beta SARS-CoV-2 vaccine in adult participants. Approximately 120 participants were screened and participants who received two or three prior doses of MVC-COV1901 were respectively placed into Group A or Group B. Eligible participants were healthy adults aged from 18 (inclusive) to 55 years who have completed two or three doses of MVC-COV1901 vaccination, with 1st and 2nd doses within 12 weeks, 2nd and 3rd doses between 12 to 24 weeks (for Group B only), and with the latest dose at least 84 days before randomization, and did not receive any other investigational or approved COVID-19 vaccines.

The trial protocol and informed consent form were approved by the Taiwan Food and Drug Administration and the institutional review boards of the Taipei Medical University Hospital (Taipei, Taiwan) and the Tri-Service General Hospital (Taipei, Taiwan). The trial was conducted in compliance with the principles of the Declaration of Helsinki and Good Clinical Practice. Safety data was monitored by an independent data and safety monitoring board (DSMB). This trial was registered at ClinicalTrials.gov: NCT05216601.

### Randomisation and masking

The randomization of each group was stratified based on site to three treatment arms, 15 mcg of MVC-COV1901, or 15 mcg or 25 mcg of MVC-COV1901-Beta in a 1:1:1 ratio. Blinding was not performed as this was an open-labeled study.

### Procedure and outcomes

The investigative product MVC-COV1901 contained 15 mcg of SARS-CoV-2 S-2P protein adjuvanted with CpG 1018 750 mcg and aluminium hydroxide 375 mcg, while MVC-COV1901-Beta contained either 15 or 25 mcg of SARS-CoV-2 Beta variant S-2P protein adjuvanted with 750 mcg CpG 1018 and 375 mcg aluminium hydroxide. Booster doses were administered as intramuscular injections of 0.5 mL of the vaccine in the deltoid region of the non-dominant arm, identical to the administration of the original MVC-COV1901 vaccine as primary series [9].

The primary safety endpoint of this study was the incidence of adverse events within 28 days of the booster administration. The primary immunogenicity endpoint was the levels of neutralising antibody titres at Visit 5 (4 weeks after the booster dose) and anti-spike immunoglobulin G (IgG) antibody titres at Visits 4 (2 weeks after the booster dose) and 5. Safety was assessed by incidences of solicited AEs for up to seven days after each vaccination and unsolicited AEs for up to 28 days after each vaccination. Other AEs, such as serious adverse events (SAEs) and adverse events of special interest (AESI), were recorded within the study period. Immunogenicity was assessed by neutralising assay with the original (WT) SARS-CoV-2 and Beta variant and IgG titres in terms of geometric mean titre (GMT) and GMT ratio. Pseudovirus neutralising assays with the Omicron variant (BA.4/BA.5 subvariant) pseudovirus were performed with samples from Visits 2 (baseline) and 5.

Neutralising antibody titres against live SARS-CoV-2 virus were performed with original SARS-CoV-2 (hCoV-19/Taiwan/4/2020, GISAID EPI_ISL_411927) and Beta variant (B.1.351, hCoVC19/Taiwan/1013) [11]. Briefly, two-fold serial dilutions of serum samples were mixed with equal volumes of 100 TCID_50_/50 μL of virus and incubated at 37 °C for one hour. After incubation, the mixtures were added to Vero E6 cells and incubated at 37 °C for four to five days. The neutralising antibody titre was defined as the reciprocal of the highest dilution capable of neutralising 50% of the infection (NT_50_) calculated using the Reed-Muench method. Anti-SARS-CoV-2 spike immunoglobulin (IgG) levels were measured by enzyme-linked immunosorbent assay (ELISA) using custom-made 96-well plates coated with S-2P antigen [10].

Pseudotyped lentivirus with spike proteins of Wuhan wildtype or Omicron (BA.4/BA.5, both possess identical spike protein sequences) variant was used in pseudovirus neutralisation assay conducted as reported previously [8]. HEK-293-hAce2 cells were inoculated with two-fold serial dilutions of serum samples mixed with equal volumes of pseudovirus. Fifty percent inhibition dilution titres (ID_50_) were calculated with uninfected cells as 100% neutralisation and cells transduced with the virus as 0% neutralisation. The mutations for the Omicron variant (BA.4/BA.5) used in the spike sequence for pseudovirus construction were derived from the WHO source: T19I, del24-26, A27S, del69-70, G142D, V213G, G339D, S371F, S373P, S375F, T376A, D405N, R408S, K417N, N440K, L452R, S477N, T478K, E484A, F486V, Q498R, N501Y, Y505H, D614G, H655Y, N679K, P681H, N764K, D796Y, Q954H, N969K [3].

Frozen peripheral blood mononuclear cells were thawed and used to set up the memory B cell (MBC) assay. Peripheral blood mononuclear cell (PBMC) suspension in RPMI 1640-10% (v/v) fecal calf serum was incubated with an equal volume of polyclonal mitogens mixture, pansorbin cells (1/2500)(heat-killed, formalin-fixed Staphylococcus aureus, Sigma-Aldrich, USA), ODN 2006 (5 mcg/mL)(Class B CpG oligonucleotide, InvivoGen, USA), and pokeweed mitogen (40 ng/mL)(Lectin from *Phytolacca americana*, Sigma-Aldrich, USA), at 37°C, 5% CO2 incubator for 6 days. After incubation, cultured PBMCs were harvested, washed and resuspended in RPMI 1640-10% (v/v) fecal calf serum for following enzyme-linked immunospot assay.

For the enzyme-linked immunospot (ELISpot) assay, 96-well PVDF membrane, 96-well multiscreen filter plate (Millipore, USA) was coated with SARS-CoV-2 spike-2P wild type, SARS-CoV-2 spike-2P Beta variant or SARS-CoV-2 spike-2P Omicron variant, at 4°C, overnight. The wells coated with PBS were negative controls and those coated with goat anti-human Ig antibody (Sigma-Aldrich, USA) were used to detect all IgG, IgM and IgA-secreting B cells. The coated plate was washed with PBS and blocked with RPMI 1640-10% (v/v) fecal calf serum at 37°C for 2 hours. After washing, cultured PBMC suspension was added to wells (200,000 cultured PBMCs for spike- or PBS-coated well and 5,000 cultured PBMCs for anti-human Ig-coated well) and incubated at 37°C, 5% CO2 incubator for 16 to 18 hours. After washing, the plate was incubated with goat anti-human IgG, IgM or IgA conjugated with alkaline phosphatase (Calbiochem, USA) at 37°C for 1 hour. After washing, the plate was developed with an alkaline phosphatase conjugate substrate kit (Bio-Rad, USA) at room temperature. Spot-forming cells were detected and measured with an automatic ELISpot reader.

T cell cytokine assay was performed as described previously using ELISpot assays specific for IFN-g and IL-4 [12]. Thawed and rested PBMCs were dispensed at 1 × 10^5^ cells per well for IFN-g ELISpot assay (Human IFN-g ELISpot Kit, MABTECH) or 2 × 10^5^ cells per well for IL-4 ELISpot assay (Human IL-4 ELISpot Kit, MABTECH). Cells were stimulated with a pool of peptides derived from ancestral Wuhan strain (PepTivator SARS-CoV-2 Prot_S WT Reference Pool, Miltenyi Biotech, Germany) and Beta variant SARS-CoV-2 (PepTivator SARS-CoV-2 Prot_S B.1.351 Mutation Pool, Miltenyi Biotech, Germany) and incubated at 37 °C for 24 to 48 h. CD3-2 monoclonal antibody were used as the positive control for stimulation. Spots were counted using the CTL automatic ELISpot reader S6 Macro Analyzer (ImmunoSpot, USA). Results were expressed as spot forming units (SFU) per million PBMC.

## Statistical analysis

As this was an exploratory phase 1 clinical study, the sample size was arbitrarily determined and was not derived from a statistical estimation method and a statistical hypothesis was not used for sample size calculation in this study. All results are presented using descriptive statistics. GMT, GMT ratio and corresponding CI are calculated using an ANCOVA model with baseline log-titres, BMI (<30 or >=30 kg/m2) and comorbidity (yes or no) and sex (male or female) as a covariate. GMT ratio is defined as the geometric mean of fold increase of post-study intervention titres over the baseline titres. Prism 6.01 (GraphPad) was used for statistical analysis. Kruskal-Wallis with corrected Dunn’s multiple comparisons test was used for comparison of means of the non-parametric dataset (Figures 3 and 5), while the Mann-Whitney test was used to compare MBC frequencies at two-time points in Figures 5B and C. Linear regression was used to model the relationship between neutralization titre and IgG MBC frequency in Figure 5D

The following groups were used for the study analysis: Safety Set included all randomized participants who received the study intervention; the Full analysis set (FAS) included all randomized participants who received the study intervention, irrespective of their protocol adherence and continued participation in the study. Pre protocol set (PPS) included all participants in the FAS who receive the planned dose of randomized study intervention, and up until Visit 5, did not have laboratory-confirmed COVID-19 infection, were negative for SARS-CoV-2 anti-N tests, and did not have major protocol deviation that was judged to impact the critical immunogenicity data.

## Results

Between May and July 2022, a total of 129 adult participants were screened and 107 eligible participants were split into groups of 45 and 62 for Groups A and B, respectively (Figure 1). In terms of the demographics of the participants, all groups had similar mean age and BMI levels, although the gender ratios were less equal among the groups (Table 1). The mean intervals between the last dose of MVC-COV1901 and the booster dose were longer in Group A (223.3 to 294.5 days) than in Group B (120.9 to 128.0 days). Solicited adverse events (SAEs) are summarized in Figure 2 and tabulated in Tables S1 and S2. Unsolicited AEs are summarized in Table S3. No SAE (grade 3 AEs or higher) or AESI related to the vaccine have been reported after the booster dose. The most common solicited local and systemic effects after any booster dose were pain/tenderness (60.0∼73.3% in Group A and 57.1∼70.0% in Group B) and malaise/fatigue (33.3∼53.3% in Group A and 28.6∼40.0% in Group B), respectively. While erythema/redness (two participants in Group A) and fever (two participants in Group A and one participant in Group B) were the least common AEs. The safety profile and incidences of AEs were comparable in both groups (Figure 2, Tables S1-S3).

**Figure 1.**
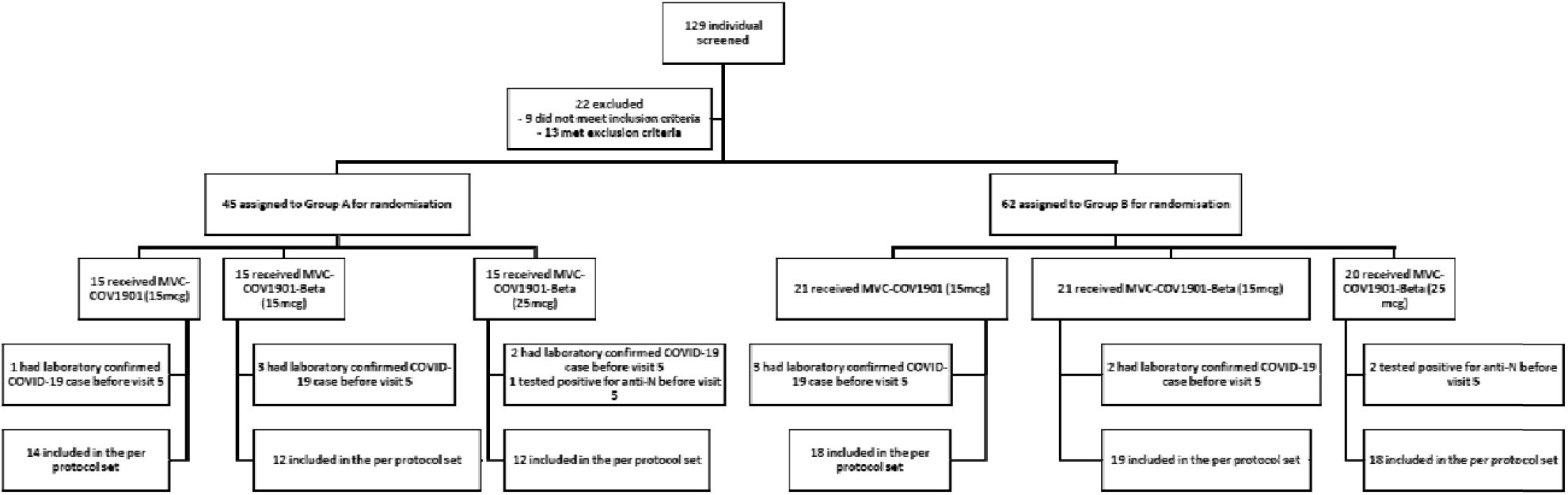
CONSORT flow diagram for the study.

**Figure 2.**
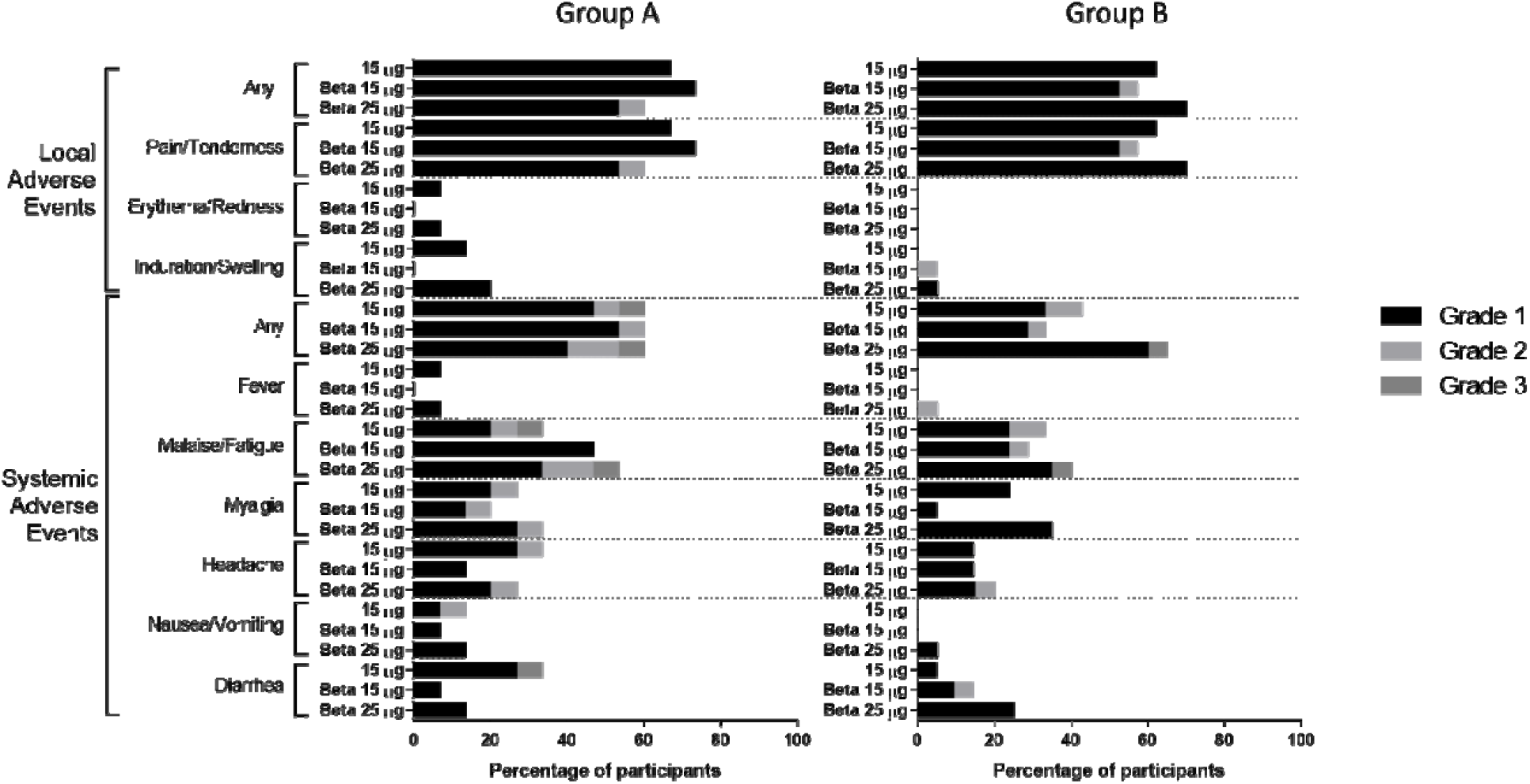
Solicited local and systemic adverse events for each of the treatment groups in Groups A and B

**Table 1.** Demographics and baseline characteristics of the participants for Group A and Group B.

|  | Group A (N=45) |  |  |
| --- | --- | --- | --- |
|  | MVC-COV1901<br>(15 mcg) | MVC-COV1901<br>(15 mcg, beta) | MVC-COV1901<br>(25 mcg, beta) |
| n | 15 | 15 | 15 |
| age mean (SD) | 36.9 (9.25) | 34.7 (7.71) | 39.5 (9.16) |
| Male | 53.3% | 73.3% | 60.0% |
| BMI | 24.991 | 24.710 | 26.215 |
| HIV-positive | 0 | 0 | 0 |
| HBsAg-positive | 0 | 0 | 0 |
| Anti-HCV antibody-positive | 0 | 0 | 0 |
| Cardiovascular disease | 0 | 0 | 0 |
| Cerebrovascular disease | 0 | 0 | 0 |
| Malignancy | 0 | 0 | 0 |
| HbA1c higher than the normal range | 0 | 1 (6.7%) | 0 |
| 1, 2 dose interval (days) mean (SD) | 33.1 (6.55) | 34.5 (4.21) | 32.6 (4.98) |
| 2, 3 dose interval (days) mean (SD) | - | - | - |
| Last dose interval (days) mean (SD) | 270.6 (101.06) | 223.3 (36.98) | 294.5 (90.88) |
|  | Group B (N=61) |  |  |
|  | MVC-COV1901<br>(15 mcg) | MVC-COV1901<br>(15 mcg, beta) | MVC-COV1901<br>(25 mcg, beta) |
| n | 21 | 21 | 20 |
| age mean (SD) | 36.8 (9.24) | 38.6 (7.53) | 38.0 (8.98) |
| Male | 38.1% | 47.6% | 55.0% |
| BMI | 24.440 | 24.210 | 24.606 |
| HIV-positive | 0 | 0 | 0 |
| HBsAg-positive | 0 | 0 | 0 |
| Anti-HCV antibody-positive | 0 | 0 | 0 |
| Cardiovascular disease | 0 | 1 (4.8%) | 0 |
| Cerebrovascular disease | 1 (4.8%) | 0 | 0 |
| Malignancy | 0 | 0 | 1 (5.0%) |
| HbA1c higher than the normal range | 2 (9.5%) | 0 | 1 (5.0%) |
| 1, 2 dose interval (days) mean (SD) | 38.7 (7.80) | 37.5 (3.04) | 37.6 (2.78) |
| 2, 3 dose interval (days) mean (SD) | 111.6 (11.82) | 113.2 (16.44) | 109.2 (9.41) |
| Last dose interval (days) mean (SD) | 120.9 (12.20) | 128.0 (28.05) | 123.2 (10.32) |

At V5, Group A participants had numerically higher levels of neutralising antibodies against the ancestral SARS-CoV-2 (WT), with the GMTs ranging from 1352.0 to 3602.8 for Group A compared to 867.9 to 1125.0 for Group B; however, the differences were not significant between the groups (Figure 3A and Table S4). Similar results were observed for the Beta variant, with the neutralizing antibody GMTs for Group A ranging from 225.6 to 1476.9 compared to 147.1 to 459.2 in Group B. In Group A, 25 mcg of MVC-COV1901-Beta significantly (p < 0.05) increased the level of neutralizing antibody against the Beta variant (1476.9 [95% CI 806.4 - 2704.8]) compared to 15 mcg of MVC-COV1901 (225.59 [128.1 – 397.2]) (Figure 3A and Table S4). However, in Group B, 15 mcg of MVC-COV1901-Beta induced the highest level of neutralizing antibody against the Beta variant (459.2 [95% CI 322.2 – 654.6]) compared to 15 mcg of MVC-COV1901 (147.1 [102.3 – 211.6) and 25 mcg of MVC-COV1901-Beta (323.8 [227.7 – 460.4]) (Figure 3A and Table S4). All participants had high levels of anti-spike IgG at Visits 4 and 5, regardless of the type of booster received or the number of prior doses of MVC-COV1901 (Figure 3B). When calculating the GMT ratio of neutralising antibody and IgG titres at V5 or V4 against the baseline (V2) titres, Group B had a minimal increase in GMT ratio compared to Group A (Figure 3C and Table S4). The increase of GMT ratio against the Beta variant was most noticeable in the 25 mcg MVC-COV1901-Beta dosage group for Group A, with V5/V2 neutralising antibody GMT ratio of 202.9 [110.8 – 371.5] and V5/V2 IgG GMT ratio of 49.4 [30.9 – 79.1] in the 25 mcg MVC-COV1901-Beta dosage group compared to for V5/V2 neutralising antibody GMT ratio of 31.0 [17.6 – 54.6] and V5/V2 IgG GMT ratio of 20.2 [13.1 – 31.1] in the 15 mcg MVC-COV1901 dosage group (Figure 3C and Table S4).

**Figure 3.**
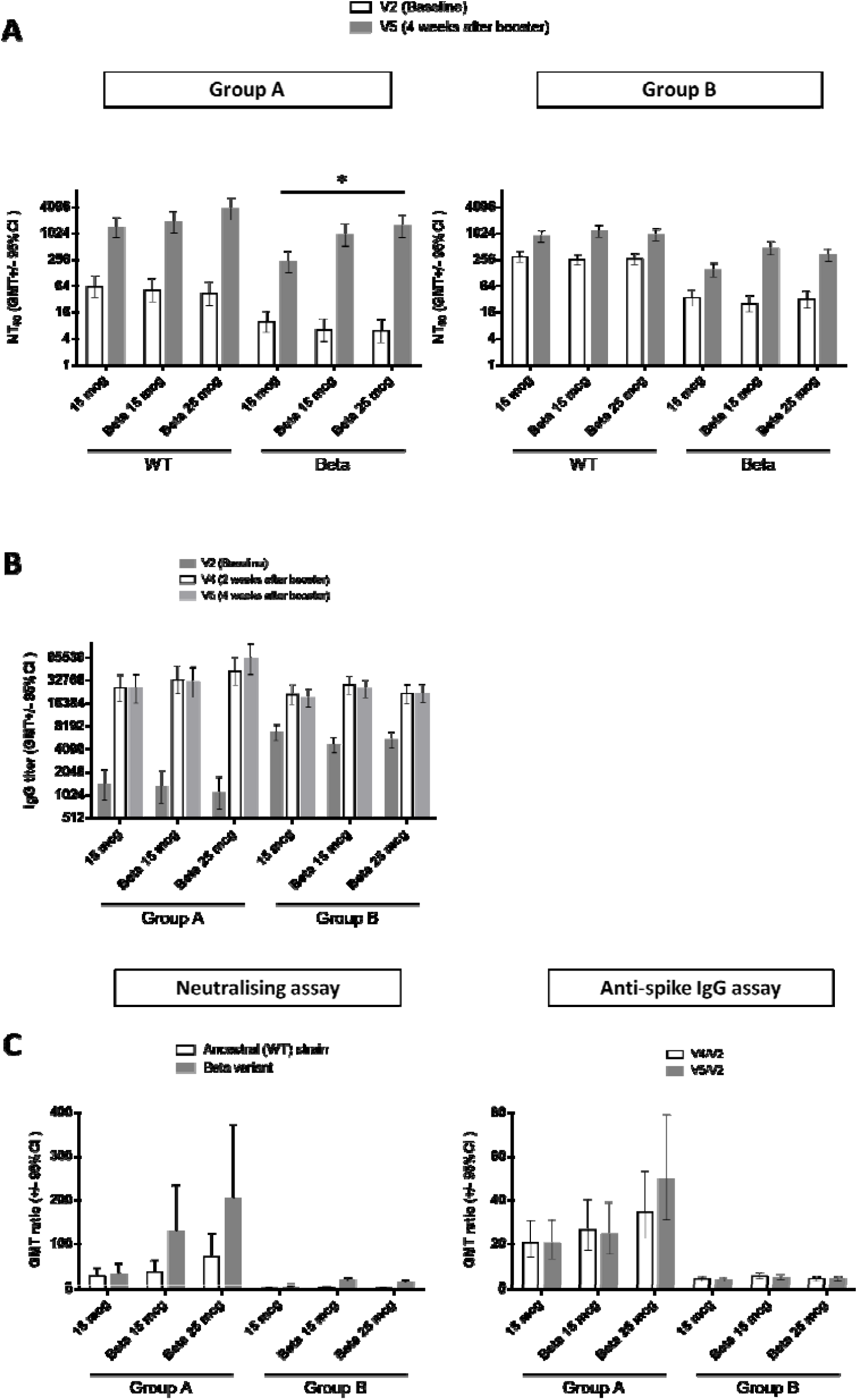
Immunogenicity of the booster dose. **A)** neutralising antibody titre against live ancestral (WT) SARS-CoV-2 and Beta variant; **B)** anti-SARS-CoV-2 spike IgG antibody titre; **C)** GMT ratio of neutralising antibody titres of V5/V2 (left) and anti-spike IgG titres at V4/V2 and V5/V2 (right). For **(A)** and **(B)**, results are expressed as symbols representing GMT and error bars represent 95% confidence intervals. For **(C)**, results are expressed as mean GMT ratio with error bars representing 95% confidence intervals. Statistical significance was calculated using the Kruskal-Wallis test with corrected Dunn’s multiple comparisons test. * = p < 0.05, ** = p < 0.01, *** = p < 0.001, **** = p < 0.0001

The immunogenicity of booster dose against the Omicron variant was investigated with the BA.4/BA.5 pseudovirus neutralization assay. In both groups at V4, all types of booster doses had uniformly high levels of neutralising antibodies against the WT pseudovirus (Figure 4). Against the BA.4/BA.5 pseudovirus, 25 mcg of MVC-COV1901-Beta elicited a significantly higher (p < 0.01) level of neutralising antibody (ID_50_ 425.7 [272.8 – 664.2]) than 15 mcg of MVC-COV1901 (ID_50_ 139.1 [76.2 – 254.1]) (Figure 4, Table S4). In Group B, this was not observed and instead, all types of booster doses resulted in similar levels of neutralising antibodies.

**Figure 4.**
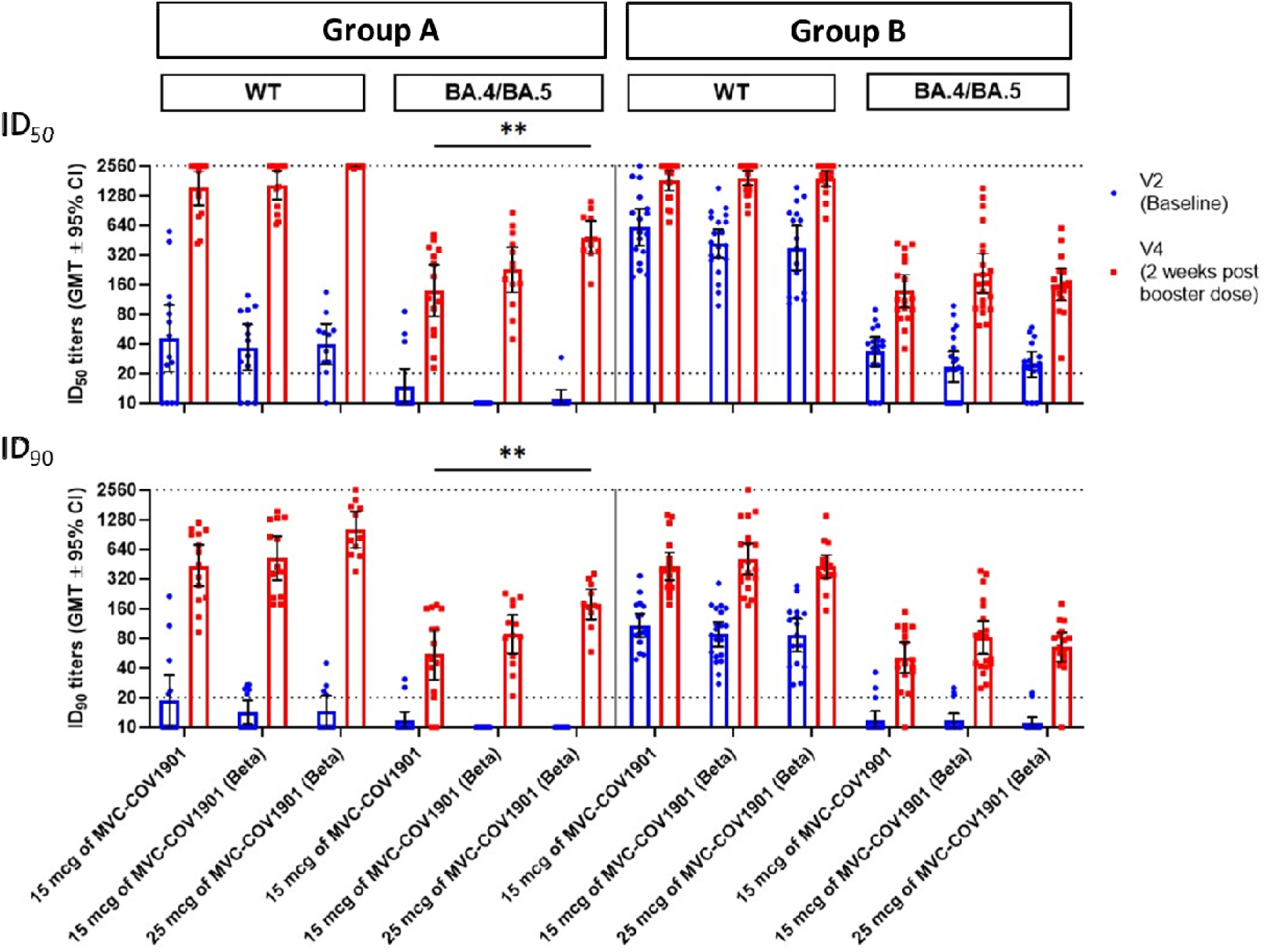
Pseudovirus neutralisation assay of pseudovirus with spike proteins of the original SARS-CoV-2 or Omicron variant (BA.4/BA.5) with serum samples from Visits 2 (baseline) and 5 (2 weeks after the booster dose). Blue and red symbols show individual titre values, while bars represent GMTs and error bars represent 95% confidence intervals. Fold differences between GMT’s are shown numerically above the upper dotted line. Dotted lines indicate the starting dilution (20; lower dotted line) and the final dilution (2560; upper dotted line) for the assay, and all values below 20 are tabulated as 10. Statistical significance was calculated using the Kruskal-Wallis test with corrected Dunn’s multiple comparisons test. * = p < 0.05, ** = p < 0.01, *** = p < 0.001, **** = p < 0.0001

Prior to the booster dose at V2, 15 of 18 (83%) Group A subjects had detectable SARS-CoV-2 spike-specific IgG memory B cells (MBCs), in which WT, Beta, or Omicron BA.1 spike-specific IgG cells accounted for around 0.5 to 0.6% of total IgG cells in the peripheral blood (Figure 5A, B; Figure S1). A total of 17 of 18 (94%) Group B subjects had detectable spike-specific IgG MBCs (Figure 5B; Figure S1). Group B subjects had significantly higher pre-existing wild type, Beta and Omicron BA.1 spike-specific IgG MBC frequencies compared to those of group A subjects (wild type, 0.6±0.1 v.s. 1.4±0.2, p = 0.002; Beta, 0.6±0.1 v.s. 1.3±0.2, p = 0.005; Omicron, 0.5±0.1 v.s. 0.9±0.1, p = 0.029, Mann-Whitney test). Pre-existing spike-specific IgM and IgA MBCs were detected in both groups as well, but there is no significant difference in the frequency between the two groups (Fig. 5B).

**Figure 5.**
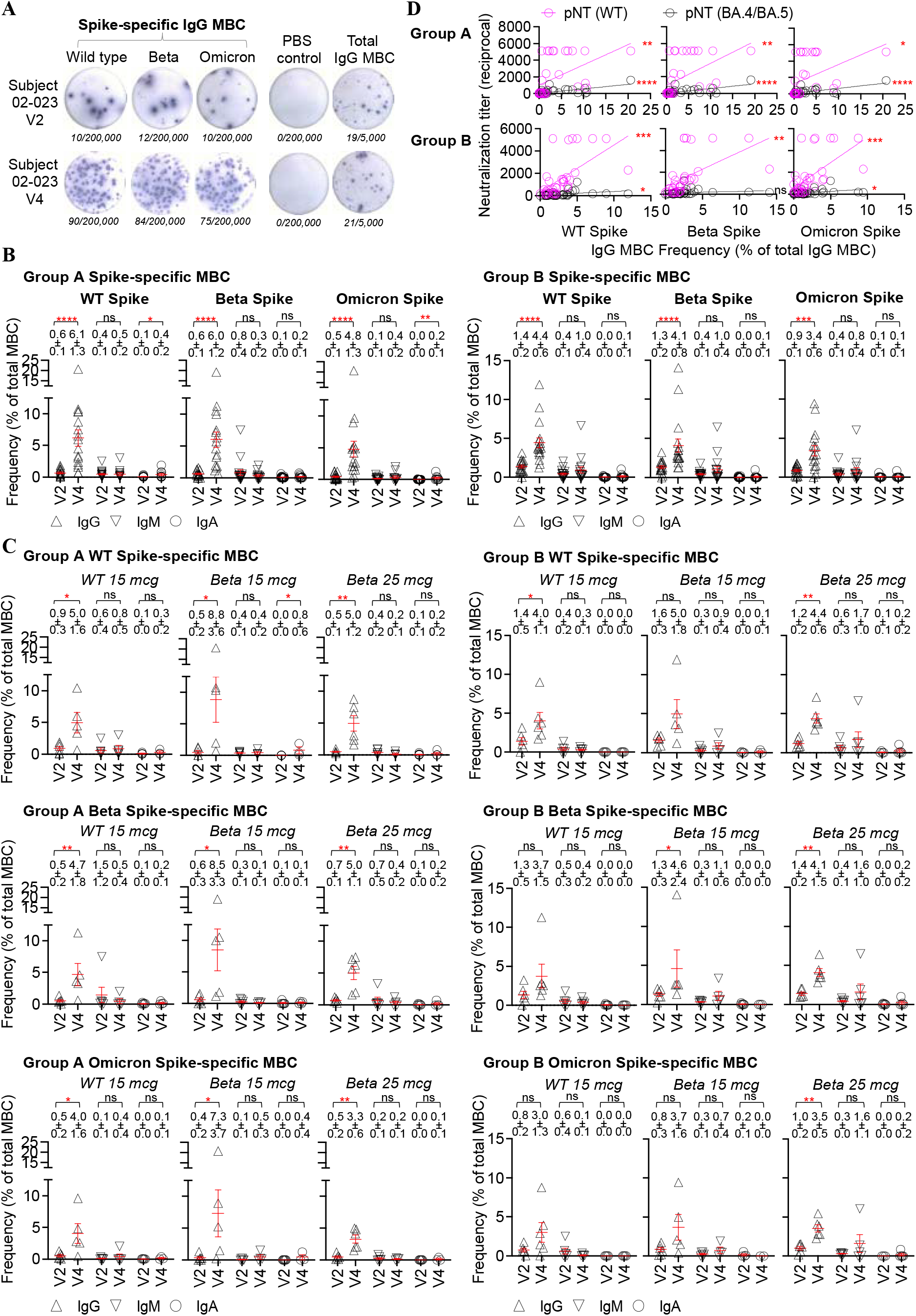
SARS-CoV-2 spike-specific memory B cell (MBC) response before and after the booster dose. (**A**) Illustration of SARS-CoV-2 spike-specific MBC frequency, measured by ELISpot. In the ELISpot assay, antigen-coated wells were used to assess antigen-specific MBC, PBS-coated wells were used as negative controls, anti-Ig coated wells were used to assess total IgG MBC. The numbers (in italics) of spots and cultured cells incubated in the ELISpot assay were shown below each image. Each spot represents an antibody-secreting cell. The frequency of antigen-specific IgG MBC was calculated as the percentage of total IgG MBC. Subject 02-023 is an adult who had two prior doses of MVC-COV1901 and received a booster dose of MVC-COV1901 containing a Beta variant spike of 15 mcg. V2, the vaccination day; V4, 14 days after the booster dose. (**B)** spike-specific MBC frequency in the peripheral blood was measured in those with two (group A) and three (group B) initial doses of MVC-COV1901, before (V2) and 14 days (V4) after the booster dose, with memory B cell ELISpot assay. Wild type (WT), Beta, and Omicron BA.1 spike-specific IgG, IgM or IgA MBC frequencies were shown in mean ± SEM in the figure. Each symbol represents a sample (subject). (**C**) spike-specific MBC frequency in the subgroups, i.e., booster dose with MVC-COV1901 containing Wuhan wild type spike, booster dose with MVC-COV1901 containing Beta variant spike 15 mcg, and booster dose with MVC-COV1901 containing Beta variant spike 25 mcg, of group A and B. Wild type (WT), Beta, and Omicron BA.1 spike-specific IgG, IgM or IgA MBC frequencies were shown in mean ± SEM in the figure. Each symbol represents a sample (subject). (**D**) Relationship of spike-specific IgG MBC frequency and serological neutralisation titre with wild type and Omicron variant BA.4/BA.5 SARS-CoV-2 pseudovirus among group A and B subjects. Linear regression was used to model the relationship between two variables. pNT, pseudovirus neutralisation titre. Mann-Whitney test was used to compare MBC frequencies at two-time points. *p < 0.05, **p < 0.01, ***p < 0.001, ****p < 0.0001. ns, not significant.

After the booster dose, a significant increase in spike-specific MBC frequency was observed in both Group A and B subjects, of which IgG MBC response dominated (Figure 5B), followed by IgM or IgA MBC responses, indicating the elicitation of immune memory to SARS-CoV-2 spike. At two weeks after the booster dose (V4), the WT, Beta, and Omicron BA.1 spike-specific IgG MBC frequency averaged 6.1±1.3, 6.0±1.2, and 4.8±1.3 of total IgG cells (P < 0.0001 for all comparisons between pre-existing and elicited responses), respectively, in Group A (Figure 5B). Enhanced spike-specific IgG MBC responses imparted by the booster were also observed in Group B at day 14 (Figure 5B). Although Group B subjects produced a relatively lower V4 wild type, Beta or Omicron BA.1 spike-specific IgG MBC frequency compared to that of Group A, the difference was not statistically significant.

All treatment groups in Group A elicited a significantly higher WT, Beta or Omicron BA.1 spike-specific IgG MBC frequency than that at the baseline (Figure 5C). Those with Beta 15 mcg booster produced a relatively higher wild type, Beta or Omicron BA.1 spike-specific IgG MBC frequency at V4 than those with wild type or Beta 25 mcg booster, but the difference was not statistically significant (Figure S2).

Within Group B, those with the original MVC-COV1901 booster produced an elevated spike-specific IgG MBC frequency at V4, but did not result in significant increases in Beta or Omicron BA.1 spike-specific MBC frequency (Figure 5C). Those with Beta 15 mcg booster produced a significantly higher Beta spike-specific IgG MBC response at V4 and those with Beta 25 mcg booster produced significantly higher wild type, Beta and Omicron BA.1 spike-specific IgG MBC frequencies at V4 that those at the baseline (Figure 5C). Nevertheless, similar WT, Beta or Omicron BA.1 spike-specific IgG MBC frequencies were detected among three subgroups at V4 (Figure S2).

Significant correlations between spike-specific MBC frequency and serological neutralisation titre were observed for Group A and B subjects, indicating a potential role of spike-specific B cell response in the development of antibody immunity upon SARS-CoV-2 immunization (Figure 5D; Figure S3).

T cell immune response was investigated by the production of interferon-gamma (IFN-γ) and IL-4 for Th-1 and Th-2 responses, respectively. Results indicated a generally Th-1-biased T cell response based on a higher amount of IFN-g induction than IL-4 induction, particularly in Group A (Figure S4).

## Discussion

This was the first clinical trial to investigate the safety and immunogenicity of a Beta variant version of the CpG 1018-adjuvanted subunit SARS-CoV-2 vaccine, MVC-COV1901-Beta, for its use as a heterologous booster dose following two or three doses of MVC-COV1901. The safety profile of MVC-COV1901-Beta was in line with that of the original MVC-COV1901, with pain/tenderness and malaise/fatigue as the most common adverse events while incidences of fever were rarely reported (Figure 2) [9, 11].

In this study, boosting with MVC-COV1901-Beta with 25 mcg of Beta S-2P protein has shown to significantly increase the neutralising antibody titre against the Beta variant as well as the Omicron variant pseudovirus compared to boosting with 15 mcg of MVC-COV1901 following two doses of MVC-COV1901 (Figures 3 and 4). The finding is consistent with our previous study, though, with the prototype antigen, which showed the increase of cross-reactivities against VoCs when antigen amount increases while adjuvant remained unchanged [13]. However, the levels of increase in neutralising antibody titre were lower in participants who had received three doses of MVC-COV1901 prior to boosting (Group B) (Figures 3 and 4). We attribute this to the differences in intervals between the last dose of MVC-COV1901 to the booster dose, which in Group A ranged from a mean of 223.3 to 294.5 days, while ranged from a mean of 109.2 to 128 days for Group B (Table 1). As less time has passed in Group B following the last vaccination compared to Group A, the baseline titres for Group B were higher and the boosting effect was less dramatic than that of Group A. In addition, in the results from our previous study in the course of three doses of MVC-COV1901 vaccination, the rate of neutralising antibody titre decay was slower after the third (booster) dose compared to the second dose, which could also explain the higher baseline titres in Group B [9].

Studies before the emergence of the Omicron variant demonstrated a correlation between the neutralizing antibody levels and the protection from infection [14]. However, the correlation between the neutralizing antibody level and the protection from the Omicron variant infection is yet to be determined. The vastly decreased protection against infection due to the vaccine escape strain reported from vaccine effectiveness studies indicates that none of the homologous regimens based on the prototype vaccine could be an optimal and durable tool to control the transmission and infection [1]. The revised target product profile for the COVID-19 vaccine published by WHO in April 2022 reflected the paradigm shift and emphasized the role of a booster vaccine for protection against severe outcomes, including hospitalization, and long COVID [15]. T-cell immune response against SARS-CoV-2 is known to play a crucial role in improving the breadth of coverage against VoCs, and offering protection against severe outcomes [16]. In patients with immune-mediated inflammatory diseases on B cell depleting therapies breakthrough infections were frequent and associated with severe outcomes [17]. These findings indicate that to achieve the revised target product profile of the COVID vaccine under the era of the Omicron variant, the roles of T and B cell immune responses should be looked at as a whole.

This is the first study to report Omicron-specific memory B cell activities after a Beta variant version of the CpG 1018-adjuvanted subunit vaccine SARS-CoV-2 vaccine. The cellular immunity after the booster dose was shown by the proliferation and expansion of spike-specific MBCs, indicating immune memory recall induced by the booster dose (Figure 5). Regardless of the type of booster used, higher proportions of MBC with IgGs specific to WT, Beta, or Omicron spikes were seen after the booster dose, thus MBCs recalled by the booster dose are cross-reactive against all strains tested (Figures 5B and C). The dominance of IgG MBC expansion also reflects the involvement of germinal center reaction and the establishment of spike-specific B cell pools, similar to the findings after the mRNA-1273 boost [18]. The potency and epitope recognition of spike-specific B cell repertoire elicited by subunit vaccine boost, especially those cross-reacting with the Omicron variant, require further investigation.

While homologous booster after two doses of vaccination provided higher protection against hospitalization against the Omicron variant, the vaccine efficiencies against infection still remained poor [19, 20]. To better counter the circulating variants, updated version of vaccines, including MVC-COV1901-Beta, have been developed and is in the progress of the deployment. The recently approved bivalent mRNA-1273.214 based on the ancestral and Omicron strains imparted a 5.4-fold increase in neutralising antibody response against the BA.4/BA.5 subvariants in a phase 2/3 clinical trial in participants with three prior doses of mRNA-1273 [21]. Pfizer/BioNTech have developed Omicron-adapted monovalent and bivalent versions of the vaccine which elicited a 13.5 to 19.6-fold increase and 9.1 to 10.9-fold increase, respectively, in neutralizing antibody titers against the Omicron BA.1 variant in a phase 2/3 trial given as a fourth dose [6]. Another monovalent adjuvanted subunit vaccine by Sanofi/GSK also generated higher titres of neutralising antibody against the Omicron BA.1 variant compared to other vaccines tested when used as a third dose booster [22].

One of the limitations of this study includes, as stated above, the differences in the time interval between the last and booster dose in Groups A and B, thus the two groups could not be compared directly. In cellular immunity assay, we did not perform surface staining and subpopulation gating and thus we could not distinguish between single-variant from dual/multi-variant spike-specific MBCs, as has been shown for mRNA-1273 [23]. The sample size was not sufficient to compare all the endpoints across treatment arms, particularly, the efficacy.

In this interim report, we found that heterologous booster with the Beta variant construct vaccine can generate a broad immune response that cross-react with the Omicron variant. The finding support further clinical development of this Beta variant version vaccine.

## Data Availability

Data sharing is not applicable to this study, as it is an interim analysis of an ongoing clinical trial.

## Conflict of Interest Disclosure

C. E. L., L. T.-C.- L., C.-C. W., W.-H. T., W.-C. L., and C. C. are employees of Medigen Vaccine Biologics Corporation. M.-C. L., N.-C. W., and K.-Y. A. H. declared no conflict of interest.

All authors have reviewed and agreed to the final version of the manuscript.

## Author Contributions

Concept, design, and leading the clinical trial: C. E. L. and C. C.

Acquisition and interpretation of data: M.-C. L., N.-C. W., and K.-Y. A. H.

Data analysis and Drafting of the manuscript: C. E. L., L. T.-C. L., and C. C.

Vaccine production: C.-C. W., W.-H. T., and W.-C. L.

## Acknowledgements

We thank the Institute of Biomedical Sciences of Academia Sinica for performing the live virus and pseudovirus neutralization assay.

