## Supplementary tables for "A Phase I, Prospective, Randomized, Open-labeled Study to Evaluate the Safety, Tolerability, and Immunogenicity of Booster Dose with MVC-COV1901 or MVC-COV1901(Beta) SARS-CoV-2 Vaccine in Adults"

**Table S1. Solicited Local Adverse Events after the Booster Dosing**

| | Group A: Two Prior Doses of MVC-COV1901 | | | | Group B: Three Prior Doses of MVC-COV1901 | | | |
|---|---|---|---|---|---|---|---|---|
| | **MVC-COV1901 (15 mcg) (N = 15), n (%)** | **MVC-COV1901 (15 mcg, Beta) (N = 15), n (%)** | **MVC-COV1901 (25 mcg, Beta) (N = 15), n (%)** | **All Participants (N = 45), n (%)** | **MVC-COV1901 (15 mcg) (N = 21), n (%)** | **MVC-COV1901 (15 mcg, Beta) (N = 21), n (%)** | **MVC-COV1901 (25 mcg, Beta) (N = 20), n (%)** | **All Participants (N = 62), n (%)** |
| **Any Solicited Local AEs** | 10 (66.7) | 11 (73.3) | 9 (60.0) | 30 (66.7) | 13 (61.9) | 12 (57.1) | 14 (70.0) | 39 (62.9) |
| Grade 1 | 10 (66.7) | 11 (73.3) | 8 (53.3) | 29 (64.4) | 13 (61.9) | 11 (52.4) | 14 (70.0) | 38 (61.3) |
| Grade 2 | 0 | 0 | 1 (6.7) | 1 (2.2) | 0 | 1 (4.8) | 0 | 1 (1.6) |
| Grade 3 | 0 | 0 | 0 | 0 | 0 | 0 | 0 | 0 |
| **Pain/Tenderness** | 10 (66.7) | 11 (73.3) | 9 (60.0) | 30 (66.7) | 13 (61.9) | 12 (57.1) | 14 (70.0) | 39 (62.9) |
| Grade 1 | 10 (66.7) | 11 (73.3) | 8 (53.3) | 29 (64.4) | 13 (61.9) | 11 (52.4) | 14 (70.0) | 38 (61.3) |
| Grade 2 | 0 | 0 | 1 (6.7) | 1 (2.2) | 0 | 1 (4.8) | 0 | 1 (1.6) |
| Grade 3 | 0 | 0 | 0 | 0 | 0 | 0 | 0 | 0 |
| **Erythema/Redness** | 1 (6.7) | 0 | 1 (6.7) | 2 (4.4) | 0 | 0 | 0 | 0 |
| Grade 1 | 1 (6.7) | 0 | 1 (6.7) | 2 (4.4) | 0 | 0 | 0 | 0 |
| Grade 2 | 0 | 0 | 0 | 0 | 0 | 0 | 0 | 0 |
| Grade 3 | 0 | 0 | 0 | 0 | 0 | 0 | 0 | 0 |
| **Induration/Swelling** | 2 (13.3) | 0 | 3 (20.0) | 5 (11.1) | 0 | 1 (4.8) | 1 (5.0) | 2 (3.2) |
| Grade 1 | 2 (13.3) | 0 | 3 (20.0) | 5 (11.1) | 0 | 0 | 1 (5.0) | 1 (1.6) |
| Grade 2 | 0 | 0 | 0 | 0 | 0 | 1 (4.8) | 0 | 1 (1.6) |
| Grade 3 | 0 | 0 | 0 | 0 | 0 | 0 | 0 | 0 |

Abbreviations: N = number of subjects in the population; n = number of subjects in the specific category. % = percentage of subjects with N as the denominator.

**Table S2. Solicited Systemic Adverse Events after the Booster Dosing**

| | Group A: Two Prior Doses of MVC-COV1901 | | | | Group B: Three Prior Doses of MVC-COV1901 | | | |
|---|---|---|---|---|---|---|---|---|
| | MVC-COV1901 (15 mcg) (N = 15), n (%) | MVC-COV1901 (15 mcg, Beta) (N = 15), n (%) | MVC-COV1901 (25 mcg, Beta) (N = 15), n (%) | All Participants (N = 45), n (%) | MVC-COV1901 (15 mcg) (N = 21), n (%) | MVC-COV1901 (15 mcg, Beta) (N = 21), n (%) | MVC-COV1901 (25 mcg, Beta) (N = 20), n (%) | All Participants (N = 62), n (%) |
| **Any Solicited Systemic AEs** | 9 (60.0) | 9 (60.0) | 9 (60.0) | 27 (60.0) | 9 (42.9) | 7 (33.3) | 13 (65.0) | 29 (46.8) |
| Grade 1 | 7 (46.7) | 8 (53.3) | 6 (40.0) | 21 (46.7) | 7 (33.3) | 6 (28.6) | 12 (60.0) | 25 (40.3) |
| Grade 2 | 1 (6.7) | 1 (6.7) | 2 (13.3) | 4 (8.9) | 2 (9.5) | 1 (4.8) | 0 | 3 (4.8) |
| Grade 3 | 1 (6.7) | 0 | 1 (6.7) | 2 (4.4) | 0 | 0 | 1 (5.0) | 1 (1.6) |
| **Fever** | 1 (6.7) | 0 | 1 (6.7) | 2 (4.4) | 0 | 0 | 1 (5.0) | 1 (1.6) |
| Grade 1 | 1 (6.7) | 0 | 1 (6.7) | 2 (4.4) | 0 | 0 | 0 | 0 |
| Grade 2 | 0 | 0 | 0 | 0 | 0 | 0 | 1 (5.0) | 1 (1.6) |
| Grade 3 | 0 | 0 | 0 | 0 | 0 | 0 | 0 | 0 |
| **Malaise/Fatigue** | 5 (33.3) | 7 (46.7) | 8 (53.3) | 20 (44.4) | 7 (33.3) | 6 (28.6) | 8 (40.0) | 21 (33.9) |
| Grade 1 | 3 (20.0) | 7 (46.7) | 5 (33.3) | 15 (33.3) | 5 (23.8) | 5 (23.8) | 7 (35.0) | 17 (27.4) |
| Grade 2 | 1 (6.7) | 0 | 2 (13.3) | 3 (6.7) | 2 (9.5) | 1 (4.8) | 0 | 3 (4.8) |
| Grade 3 | 1 (6.7) | 0 | 1 (6.7) | 2 (4.4) | 0 | 0 | 1 (5.0) | 1 (1.6) |
| **Myalgia** | 4 (26.7) | 3 (20.0) | 5 (33.3) | 12 (26.7) | 5 (23.8) | 1 (4.8) | 7 (35.0) | 13 (21.0) |
| Grade 1 | 3 (20.0) | 2 (13.3) | 4 (26.7) | 9 (20.0) | 5 (23.8) | 1 (4.8) | 7 (35.0) | 13 (21.0) |
| Grade 2 | 1 (6.7) | 1 (6.7) | 1 (6.7) | 3 (6.7) | 0 | 0 | 0 | 0 |
| Grade 3 | 0 | 0 | 0 | 0 | 0 | 0 | 0 | 0 |
| **Headache** | 5 (33.3) | 2 (13.3) | 4 (26.7) | 11 (24.4) | 3 (14.3) | 3 (14.3) | 4 (20.0) | 10 (16.1) |
| Grade 1 | 4 (26.7) | 2 (13.3) | 3 (20.0) | 9 (20.0) | 3 (14.3) | 3 (14.3) | 3 (15.0) | 9 (14.5) |

| | Group A: Two Prior Doses of MVC-COV1901 | | | | Group B: Three Prior Doses of MVC-COV1901 | | | |
|---|---|---|---|---|---|---|---|---|
| | **MVC-COV1901 (15 mcg) (N = 15), n (%)** | **MVC-COV1901 (15 mcg, Beta) (N = 15), n (%)** | **MVC-COV1901 (25 mcg, Beta) (N = 15), n (%)** | **All Participants (N = 45), n (%)** | **MVC-COV1901 (15 mcg) (N = 21), n (%)** | **MVC-COV1901 (15 mcg, Beta) (N = 21), n (%)** | **MVC-COV1901 (25 mcg, Beta) (N = 20), n (%)** | **All Participants (N = 62), n (%)** |
| Grade 2 | 1 (6.7) | 0 | 1 (6.7) | 2 (4.4) | 0 | 0 | 1 (5.0) | 1 (1.6) |
| Grade 3 | 0 | 0 | 0 | 0 | 0 | 0 | 0 | 0 |
| **Nausea/Vomiting** | 2 (13.3) | 1 (6.7) | 2 (13.3) | 5 (11.1) | 0 | 0 | 1 (5.0) | 1 (1.6) |
| Grade 1 | 1 (6.7) | 1 (6.7) | 2 (13.3) | 4 (8.9) | 0 | 0 | 1 (5.0) | 1 (1.6) |
| Grade 2 | 1 (6.7) | 0 | 0 | 1 (2.2) | 0 | 0 | 0 | 0 |
| Grade 3 | 0 | 0 | 0 | 0 | 0 | 0 | 0 | 0 |
| **Diarrhea** | 5 (33.3) | 1 (6.7) | 2 (13.3) | 8 (17.8) | 1 (4.8) | 3 (14.3) | 5 (25.0) | 9 (14.5) |
| Grade 1 | 4 (26.7) | 1 (6.7) | 2 (13.3) | 7 (15.6) | 1 (4.8) | 2 (9.5) | 5 (25.0) | 8 (12.9) |
| Grade 2 | 0 | 0 | 0 | 0 | 0 | 1 (4.8) | 0 | 1 (1.6) |
| Grade 3 | 1 (6.7) | 0 | 0 | 1 (2.2) | 0 | 0 | 0 | 0 |

Abbreviations: N = number of subjects in the population; n = number of subjects in the specific category. % = percentage of subjects with N as the denominator.

**Table S3. Summary of Unsolicited Adverse Events and Other Adverse Events**

| | Group A: Two Prior Doses of MVC-COV1901 | | | | Group B: Three Prior Doses of MVC-COV1901 | | | |
|---|---|---|---|---|---|---|---|---|
| | MVC-COV1901 (15 mcg) (N = 15), n (%) | MVC-COV1901 (15 mcg, Beta) (N = 15), n (%) | MVC-COV1901 (25 mcg, Beta) (N = 15), n (%) | All Participants (N = 45), n (%) | MVC-COV1901 (15 mcg) (N = 21), n (%) | MVC-COV1901 (15 mcg, Beta) (N = 21), n (%) | MVC-COV1901 (25 mcg, Beta) (N = 20), n (%) | All Participants (N = 62), n (%) |
| Unsolicited AEs | 4 (26.7) | 6 (40.0) | 5 (33.3) | 15 (33.3) | 6 (28.6) | 8 (38.1) | 3 (15.0) | 17 (27.4) |
| Related Unsolicited AEs | 1 (6.7) | 1 (6.7) | 2 (13.3) | 4 (8.9) | 2 (9.5) | 2 (9.5) | 2 (10.0) | 6 (9.7) |
| Unsolicited AEs ≥ Grade 3 | 0 | 0 | 0 | 0 | 0 | 0 | 0 | 0 |
| Related Unsolicited AEs ≥ Grade 3 | 0 | 0 | 0 | 0 | 0 | 0 | 0 | 0 |
| AEs ≥ Grade 3 | 0 | 0 | 0 | 0 | 0 | 0 | 0 | 0 |
| Related AEs ≥ Grade 3 | 0 | 0 | 0 | 0 | 0 | 0 | 0 | 0 |
| SAEs | 0 | 0 | 0 | 0 | 0 | 0 | 0 | 0 |
| Related SAEs | 0 | 0 | 0 | 0 | 0 | 0 | 0 | 0 |
| AESI (including pIMD) | 0 | 0 | 0 | 0 | 0 | 0 | 0 | 0 |
| VAED | 0 | 0 | 0 | 0 | 0 | 0 | 0 | 0 |
| AEs Leading to Intervention Discontinuation | 0 | 0 | 0 | 0 | 0 | 0 | 0 | 0 |
| AEs Leading to Study Withdrawal | 0 | 0 | 0 | 0 | 0 | 0 | 0 | 0 |
| Death | 0 | 0 | 0 | 0 | 0 | 0 | 0 | 0 |

Abbreviations: N = number of subjects in the population; n = number of subjects with events; AE = Adverse Event; SAE = Serious Adverse Event; AESI = Adverse events of special interest; pIMD = potential immune mediated disorders; VAED = Vaccine-associated enhanced disease.

**Table S4. Summary of immunogenicity data for Groups A and B**

| | | Group A | | | | | |
|---|---|---|---|---|---|---|---|
| | | **MVC-COV1901 (15 mcg) (n=14)** | | **MVC-COV1901-Beta (15 mcg) (n=12)** | | **MVC-COV1901-Beta (25 mcg) (n=12)** | |
| | | **[GMT]** | **95% CI** | **[GMT]** | **95% CI** | **[GMT]** | **95% CI** |
| **WT ($NT_{50}$)*** | V2 (Day 1) | **61.28** | (34.48, 108.91) | **50.74** | (27.26, 94.44) | **42.47** | (22.82, 79.03) |
| | V5 (Day 29) | **1352.00** | (797.38, 2292.40) | **1805.02** | (1023.61, 3182.94) | **3602.75** | (2036.68, 6373.05) |
| **WT GMT ratio**** | V5/V2 | **26.29** | (15.51, 44.58) | **35.10** | (19.91, 61.90) | **70.06** | (39.61, 123.94) |
| **Beta ($NT_{50}$)*** | V2 (Day 1) | **9.75** | (5.57, 17.08) | **6.29** | (3.43, 11.52) | **5.99** | (3.27, 10.98) |
| | V5 (Day 29) | **225.59** | (128.13, 397.17) | **931.34** | (509.33, 1703.00) | **1476.85** | (806.39, 2704.76) |
| **Beta GMT ratio**** | V5/V2 | **30.98** | (17.60, 54.55) | **127.92** | (69.96, 233.91) | **202.85** | (110.76, 371.51) |
| **Anti-spike IgG (WT)** | V2 (Day 1) | **1384.95** | (876.59, 2188.12) | **1301.00** | (793.82, 2132.21) | **1073.76** | (655.17, 1759.79) |
| | V4 (Day 15) | **25878.60** | (17387.24, 38516.84) | **32938.41** | (21459.28, 50558.02) | **43208.61** | (28085.34, 66475.38) |
| | V5 (Day 29) | **25257.76** | (16360.67, 38993.18) | **30672.19** | (19210.70, 48971.84) | **61907.00** | (38676.11, 99091.57) |
| **IgG GMT ratio**** | V4/V2 | **20.65** | (13.88, 30.74) | **26.29** | (17.13, 40.35) | **34.48** | (22.41, 53.05) |
| | V5/V2 | **20.16** | (13.06, 31.12) | **24.48** | (15.33, 39.08) | **49.41** | (30.87, 79.08) |
| **Pseudovirus NT $ID_{50}$ (WT)** | V2 (Day 1) | **45.12** | (98.07, 20.76) | **35.97** | (19.84, 65.21) | **40.62** | (26.33, 62.66) |
| | V4 (Day 15) | **1520.96** | (1023.93, 2259.24) | **1723.40** | (1227.59, 2419.44) | **2315.39** | (1879.03, 2853.08) |
| **Pseudovirus NT $ID_{50}$ (BA.4/BA.5)** | V2 (Day 1) | **14.50** | (9.39, 22.38) | **10.00** | (10.00, 10.00) | **10.93** | (8.99, 13.30) |
| | V4 (Day 15) | **139.11** | (76.16, 254.09) | **240.10** | (137.63, 418.86) | **425.65** | (272.79, 664.17) |
| **Pseudovirus NT $ID_{90}$ (WT)** | V2 (Day 1) | **18.53** | (10.30, 33.33) | **14.58** | (10.82, 19.63) | **14.05** | (10.02, 19.68) |
| | V4 (Day 15) | **434.71** | (267.37, 706.78) | **558.18** | (328.24, 949.20) | **888.27** | (551.22, 1431.43) |
| **Pseudovirus NT $ID_{90}$ (BA.4/BA.5)** | V2 (Day 1) | **11.58** | (9.33, 14.38) | **10.00** | (10.00, 10.00) | **10.00** | (10.00, 10.00) |
| | V4 (Day 15) | **54.52** | (30.41, 97.75) | **92.95** | (57.80, 149.50) | **157.99** | (105.15, 237.39) |

| | | Group B | | | | | |
|---|---|---|---|---|---|---|---|
| | | **MVC-COV1901 (15 mcg) (n=18)** | | **MVC-COV1901-Beta (15 mcg) (n=19)** | | **MVC-COV1901-Beta (25 mcg) (n=18)** | |
| | | **[GMT]** | **95% CI** | **[GMT]** | **95% CI** | **[GMT]** | **95% CI** |
| **WT ($NT_{50}$)*** | V2 (Day 1) | **292.89** | (220.46, 389.11) | **253.89** | (192.56, 334.76) | **263.29** | (198.18, 349.79) |
| | V5 (Day 29) | **867.93** | (629.98, 1195.74) | **1124.98** | (824.29, 1535.36) | **928.54** | (680.78, 1266.48) |
| **WT GMT ratio**** | V5/V2 | **3.25** | (2.36, 4.47) | **4.21** | (3.08, 5.74) | **3.47** | (2.55, 4.74) |
| **Beta ($NT_{50}$)*** | V2 (Day 1) | **34.02** | (21.76, 53.19) | **25.09** | (16.24, 38.77) | **31.17** | (19.94, 48.74) |
| | V5 (Day 29) | **147.14** | (102.32, 211.61) | **459.23** | (322.19, 654.56) | **323.78** | (227.68, 460.43) |
| **Beta GMT ratio**** | V5/V2 | **5.03** | (3.49, 7.23) | **15.69** | (11.01, 22.36) | **11.06** | (7.78, 15.73) |
| **Anti-spike IgG (WT)** | V2 (Day 1) | **6712.17** | (5261.78, 8562.35) | **4620.77** | (3645.90, 5856.32) | **5307.28** | (4160.46, 6770.21) |
| | V4 (Day 15) | **21078.01** | (15675.17, 28343.06) | **28179.38** | (21170.75, 37508.24) | **21638.16** | (16216.34, 28872.72) |
| | V5 (Day 29) | **19014.66** | (14364.79, 25169.70) | **24953.2** | (19040.94, 32701.23) | **21889.96** | (16782.37, 28552.02) |
| **IgG GMT ratio**** | V4/V2 | **3.86** | (2.87, 5.19) | **5.16** | (3.87, 6.87) | **3.96** | (2.97, 5.28) |
| | V5/V2 | **3.48** | (2.63, 4.61) | **4.57** | (3.49, 5.99) | **4.01** | (3.07, 5.23) |
| **Pseudovirus NT $ID_{50}$ (WT)** | V2 (Day 1) | **611.33** | (400.87, 932.27) | **388.14** | (271.86, 554.15) | **408.18** | (254.72, 654.10) |
| | V4 (Day 15) | **1831.75** | (1449.69, 2314.49) | **1919.17** | (1595.53, 2308.44) | **1919.51** | (1613.19, 2284.00) |
| **Pseudovirus NT $ID_{50}$ (BA.4/BA.5)** | V2 (Day 1) | **33.28** | (23.69, 46.76) | **22.31** | (15.16, 32.83) | **25.66** | (19.46, 33.84) |
| | V4 (Day 15) | **136.83** | (93.16, 200.95) | **222.44** | (135.10, 366.22) | **154.62** | (109.03, 219.28) |
| **Pseudovirus NT $ID_{90}$ (WT)** | V2 (Day 1) | **108.97** | (82.54, 143.87) | **81.60** | (60.18, 110.66) | **92.07** | (64.51, 131.40) |
| | V4 (Day 15) | **428.68** | (308.92, 594.85) | **533.7** | (360.77, 789.53) | **417.94** | (328.33, 532.01) |
| **Pseudovirus NT $ID_{90}$ (BA.4/BA.5)** | V2 (Day 1) | **11.85** | (9.69, 14.50) | **11.46** | (9.80, 13.42) | **11.35** | (9.82, 13.12) |
| | V4 (Day 15) | **50.83** | (35.40, 72.98) | **84.52** | (55.02, 129.83) | **63.74** | (46.65, 87.10) |

*$NT_{50}$ GMT and corresponding CI are calculated using an ANCOVA model with baseline log-titers, BMI (<30 or >=30 kg/m2) and comorbidity (yes or no) and sex (male or female) as covariate.

**GMT ratio is defined as the geometric mean of fold increase of post-study intervention titres over the baseline titres
