## Supplementary material for "A Phase I, Prospective, Randomized, Open-labeled Study to Evaluate the Safety, Tolerability, and Immunogenicity of Booster Dose with MVC-COV1901 or MVC-COV1901(Beta) SARS-CoV-2 Vaccine in Adults": Supplemenary Figures

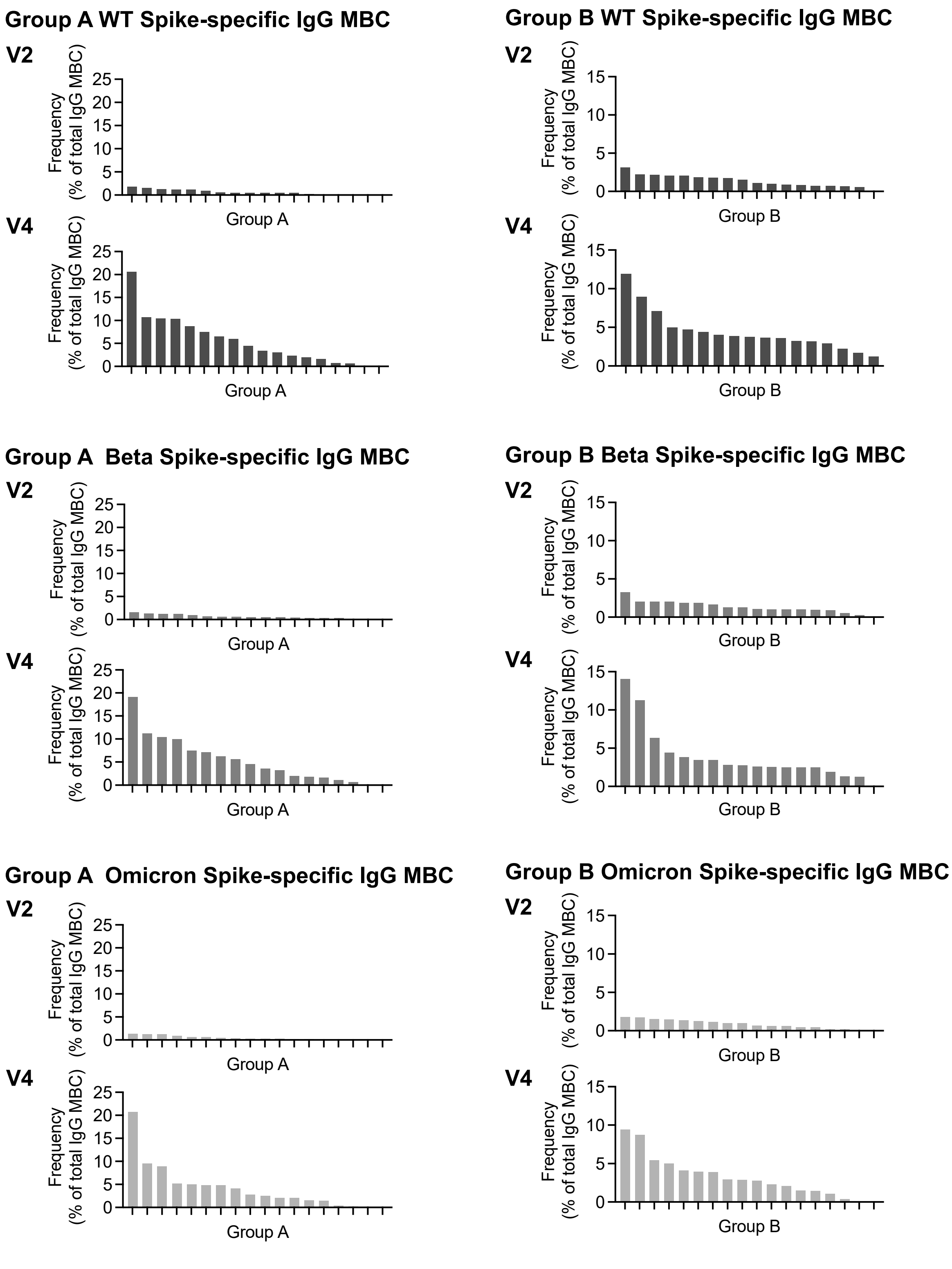


**Supplemental Figure 1. Summary data of Spike-specific IgG memory B cell frequency prior to (V2) and 14 days after (V4) the booster dose in group A and B subjects.** Those who had two and three prior MVC-COV191 belonged to group A and B, respectively. WT, wild type. MBC, memory B cell

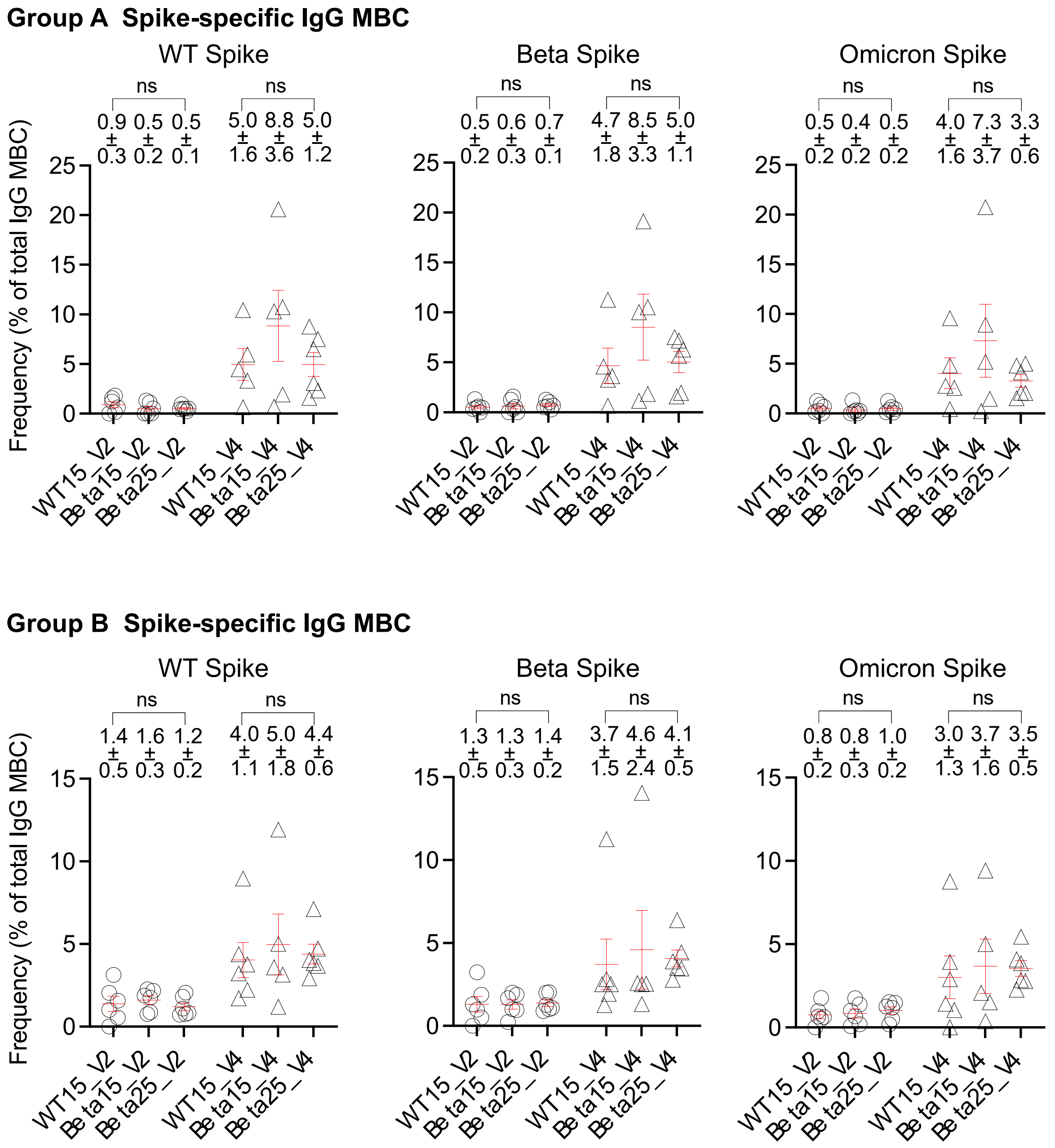


**Supplemental Figure 2. Comparison of Spike-specific IgG MBC frequencies among subgroups in group A and B subjects.** There are three subgroups, i.e., booster dose with MVC-COV1901 containing Wuhan wild type Spike, booster dose with MVC-COV1901 containing Beta variant Spike 15 µg, and booster dose with MVC-COV1901 containing Beta variant Spike 25 µg, for each of group A and B. One-way ANOVA was used to compare the difference among subgroups. ns, not significant. V2, the vaccination day; V4, 14 days after the booster dose.

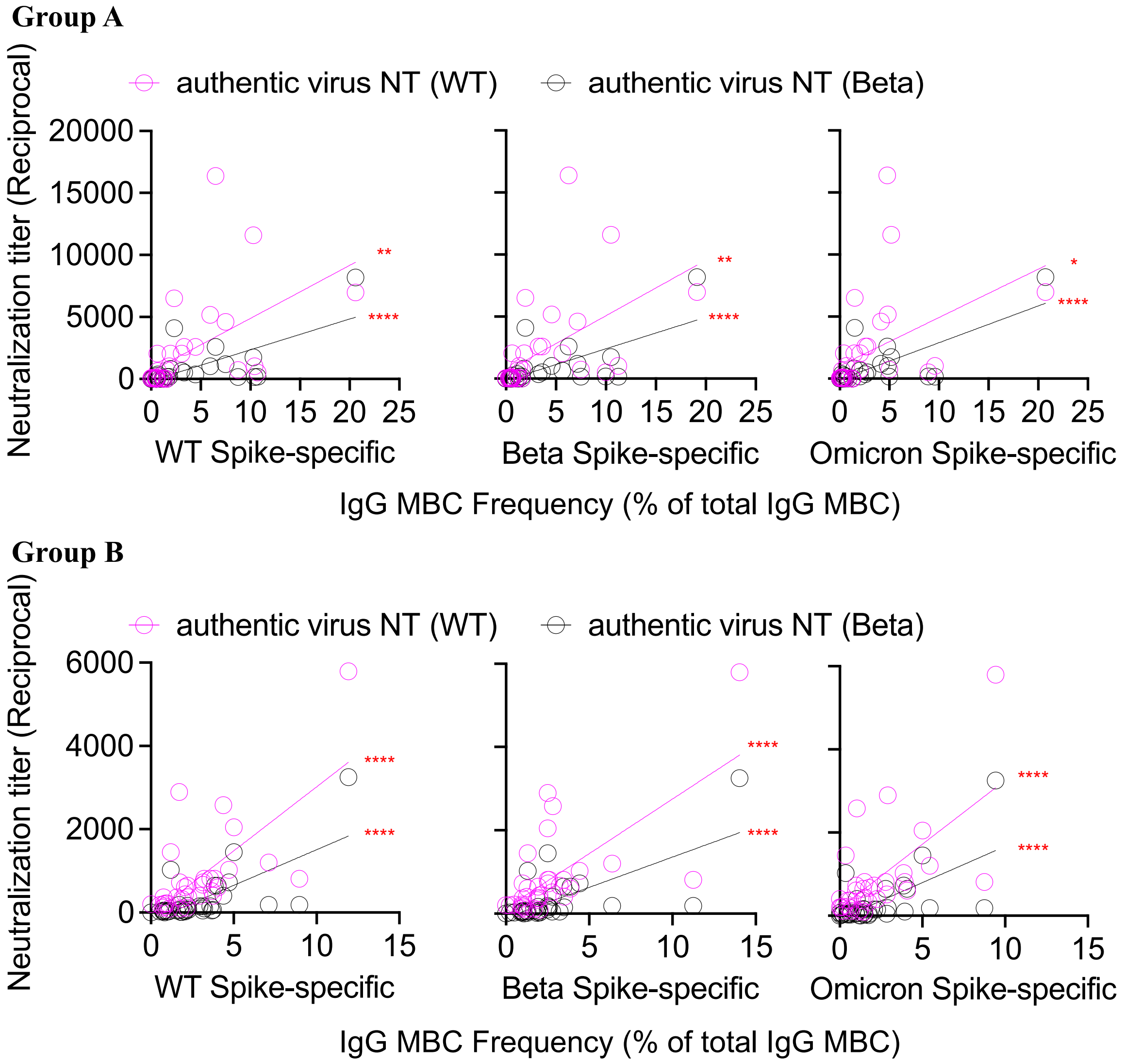


**Supplemental Figure 3. Relationship of Spike-specific IgG MBC frequency and serological neutralization titer with authentic wild type and Beta variant viruses among group A and B subjects.** Linear regression was used to model the relationship between two variables. WT, wild type; NT, neutralization test. *p < 0.05, **p < 0.01, ****p < 0.0001.

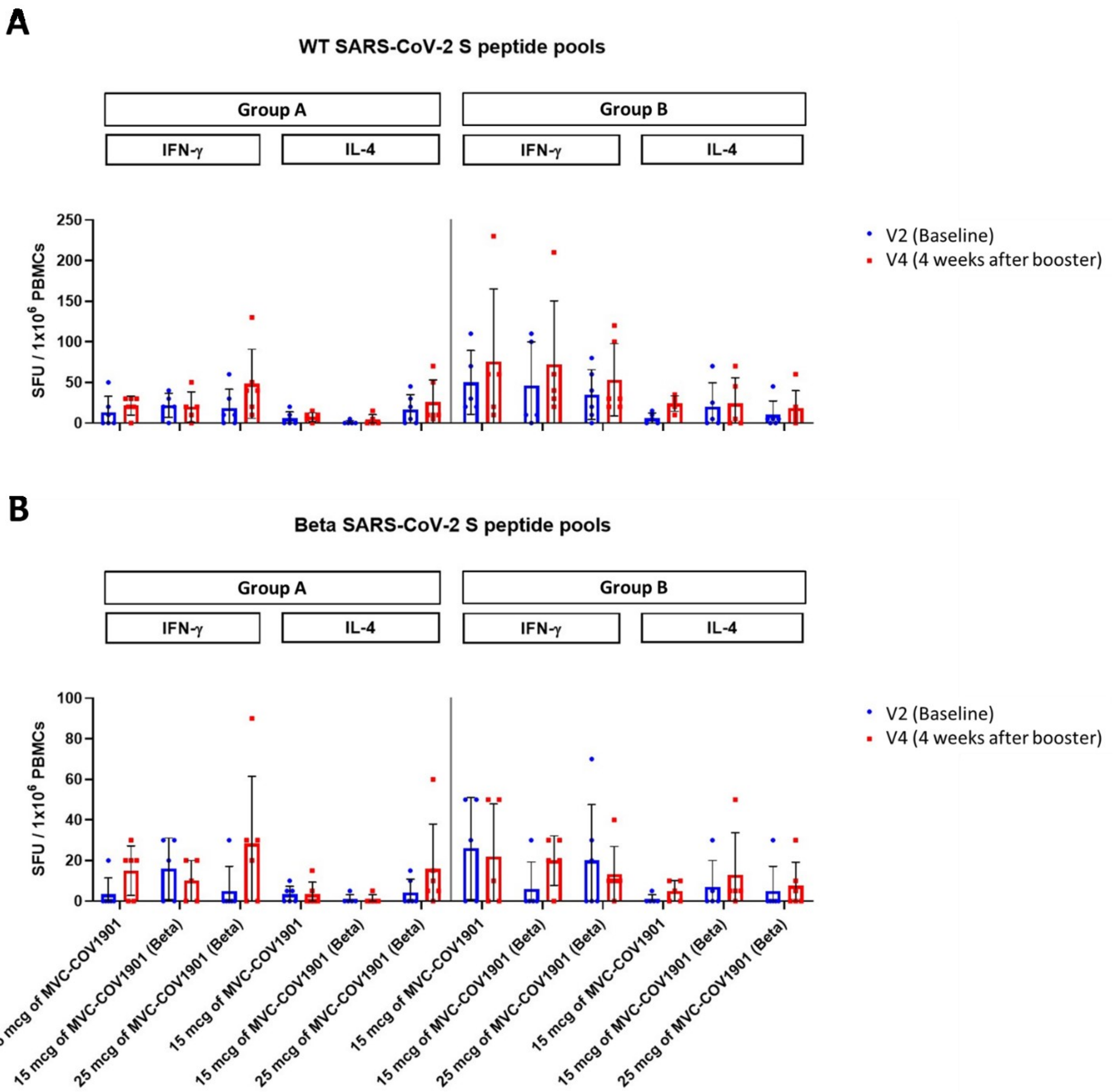


**Supplemental Figure 4. Cytokine production induced by MVC-COV1901 or MVC-COV1901-Beta boosters.** PBMC samples were stimulated with peptide pools derived from the ancestral Wuhan (WT) or Beta variant spike protein of SARS-CoV-2. The results are shown in mean spot forming units (SFU) per million of PBMCs with error bars representing standard deviations.
